# Ongoing transmission of human onchocerciasis in the district of Mont Ngafula 1 in Kinshasa after two decades of uninterrupted onchocerciasis annual mass campaigns using community directed treatment with Ivermectin strategy

**DOI:** 10.1101/2023.05.10.23289767

**Authors:** Makenga Bof Jean Claude, Mansiangi Paul, Zanga Josué, Ilunga Félicien, Ako Aime Gilles Adjami, Sanfo Moussa Sounkalo, Didier Bakajika, Coppieters Yves

**Author notes:** **Corresponding author** Makenga Bof Jean Claude, Ecole de Santé Publique, Université Libre de Bruxelles (ULB), Route de Lennik 808, 1070 Brussels, Belgium. Deceased (Mansiangi Paul).

## Abstract

**Background:** The district of Mont Ngafula I in Kinshasa, the capital city of the Democratic Republic of Congo (DRC) has been treating for onchocerciasis over the last two decades using community directed treatment with ivermectin strategy (CDTI). This study aimed to determine the transmission of onchocerciasis in blackflies after two decades of uninterrupted annual ivermectin mass campaigns using CDTI.

**Material:** Blackflies were collected at Kimwenza site in the district of Mont Ngafula 1 along Lukaya river from 1^st^ August 2019 to 31^st^ July 2020 using human landing catching techniques. Entomological indicators (biting rate, transmission potentials and infectivity rate) were calculated using O-150 Pool screening PCR technique.

**Results:** A total of 12,217 blackflies of *Simulium squamosum* species were collected during the study period. Two daily cycles of high biting were identified between 08:00 and 09:00 a.m. and 16:00 and 17:00. Low biting rates were observed between 11:00 a.m. and 13:00. The daily and annual biting rates were 774 and 22,380 bites/person (p = < 0.001). The infectivity rate was 0.09 % (95% CI: 0.04 - 0.17). The calculated annual transmission potential was 21).

**Conclusions:** The study showed an ongoing transmission of onchocerciasis in the study site despite two decades of uninterrupted ivermectin mass distribution campaigns using community directed treatment with ivermectin. There is an urgent need for alternative treatment strategies to accelerate the interruption of transmission of onchocerciasis.

**Author Summary:** Human onchocerciasis, also known as river blindness, is a neglected tropical disease targeted for elimination of transmission by the World Health Organization (WHO). Both epidemiological and entomological criteria are highly needed to confirm the interruption and elimination of transmission of onchocerciasis. Following rapid epidemiological mapping of Onchocerciasis conducted in 2001 in Kinshasa, the capital city of the Democratic Republic of Congo, the medical district of Mont Ngafula 1, Nsele and Binza Ozone were found to be mesoendemic for onchocerciasis and were treated using community directed treatment with ivermectin strategy (CDTI) for the last two decades. The aim of this study was to determine the impact of Ivermectin community mass distribution on onchocerciasis transmission in blackflies in the health district of Mont Ngafula 1 after two decades of preventive chemotherapy. A total of 12,217 blackflies were collected from 1^st^ August 2019 to 31^st^ July 2020 using human landing catching technique. Biting rate and annual potential transmission were calculated and infectivity rate determined using the O-150 PCR technique. The findings confirmed an ongoing transmission of onchocerciasis in blackflies in the health district of Mont Ngafula 1 despite two decades on uninterrupted Ivermectin mass administration. There is an urgent call for alternative treatment strategies to accelerate the interruption of transmission of onchocerciasis in the study site.

## 1. Introduction

Human onchocerciasis, also called “*river blindness*”, is caused by the filarial worm *Onchocerca volvulus*, transmitted to humans through exposure to repeated bites of infected blackflies of the genus *Simulium* that breed in fast-flowing streams and rivers [1]. This parasitic disease is one of the twenty neglected tropical diseases (NTDs) targeted for elimination by the World Health Organization (WHO) [2]. In 2021, more than 244.6 million people were identified worldwide as living in endemic settings requiring [3], Ivermectin preventive chemotherapy of which 243.7 were in the WHO African region.

The Democratic Republic of Congo (DRC) is one of the 27 countries in the WHO African region known to be endemic for onchocerciasis [1]. The disease is endemic in all the 26 administrative provinces of the country, including the city-province of Kinshasa [4]. First blackfly breeding sites in Kinshasa were discovered at the beginning of the 20^th^ century [5]. Subsequently, several studies carried out until 1945 in this focus determined the bio-ecology of the vector and specified its critical role in the transmission of the infection [5]. A 5-year vector control study was conducted from 1948 to 1952, using dichlorodiphenyltrichloroethane (DDT) with the aim to “eradicate” the vector in the Kinshasa focus [6]. Unfortunately, 10 years after this successful project the resurgence of blackflies was observed in the project area [7]. Rapid Epidemiological Mapping of Onchocerciasis (REMO) conducted by the National Onchocerciasis Control Program (NOCP) in 2001 with the financial and technical support of the former African Program for Onchocerciasis control (APOC) to determine the endemicity status of onchocerciasis in Kinshasa found three medical districts to be meso-endemic for onchocerciasis namely Binza-Ozone, Nsele and Mont Ngafula 1 and therefore were eligible for Ivermectin mass distribution using the community directed treatment with ivermectin (CDTI) strategy. CDTI strategy has been implemented in these three districts for almost two decades [6 - 11].

After almost two decades of CDTI strategy in the city province of Kinshasa, it is appropriate to determine its impact on the transmission in order to better guide future programmatic actions. The current study aims to determine the transmission of onchocerciasis in blackflies at the site in the medical district of Mont-Ngafula 1.

## 2. Materials and methods

### 2.1. Description of the study environment

Kinshasa, the capital of the DRC, is a large city (area: 9,965 Km^2^), with 24 municipalities and four administrative districts, with an estimated population of 11 million (population density: ±3,600 inhabitants/Km^2^) [12].

Kinshasa has a tropical savannah climate, equatorial in nature (hot and humid), with a rainy season lasting 8 months and a dry one for 4 months. The average annual temperature and precipitation are 25.3°C and 1,273.9 mm, respectively [12,13]. The dry season (tropical winter) which starts in June and ends in September, is characterized by rare and low rainfall and lowest temperatures [12,13]. During the rainy season (tropical summer), from October to May, the precipitation ranges between 1000 and 1500 mm [12,13], while the temperature ranges between 30-35ºC (peaks at 40°C) [12,13].

Rivers of various sizes originating from the hills drain the plains of Kinshasa and flow from south to north to the Congo River [18,20]. Lakes (smaller waterbodies than rivers), such as *Lac Ma Vallée* and *Lac Vert*, are also located in Kinshasa [18,20]. Some of these waterbodies exhibit geographical characteristics that enable rapid waterfalls favorable for the breeding of *Simulium vectors*, particularly, the Lukaya River in rural Kimwenza and Congo River in the urban part of the bay of Gombe [12,14].

The study was conducted in municipality of Mont Ngafula 1 [14]. Mont-Ngafula 1 is located in the southern region of Kinshasa, in the hilly areas of rural Kimwenza occupied by the Lukaya River. It is a recent settlement with a dominant highland plateau at an altitude of 493 meters near the Lukaya, little Waterfalls and tourist site “Lola Ya Bonobo” (**Figure 1**) [14]. The Kimwenza site was chosen due to lack of baseline and follow up onchocerciasis entomological studies and to assess the impact of uninterrupted preventive chemotherapy interventions on the transmission in blackflies. The municipality of Mont Ngafula was explored to locate the roosts of pre-imaginal forms of black flies, identify their fixation supports, and select the collection points using Enyong’s criteria [15]. The best collection points were defined as the one capture points of anthropophilic adult black flies for determining entomological indices (Kimwenza site) at 4° 27’ 33’’ South, 15° 17’ 20’’ East).

**Figure 1:**
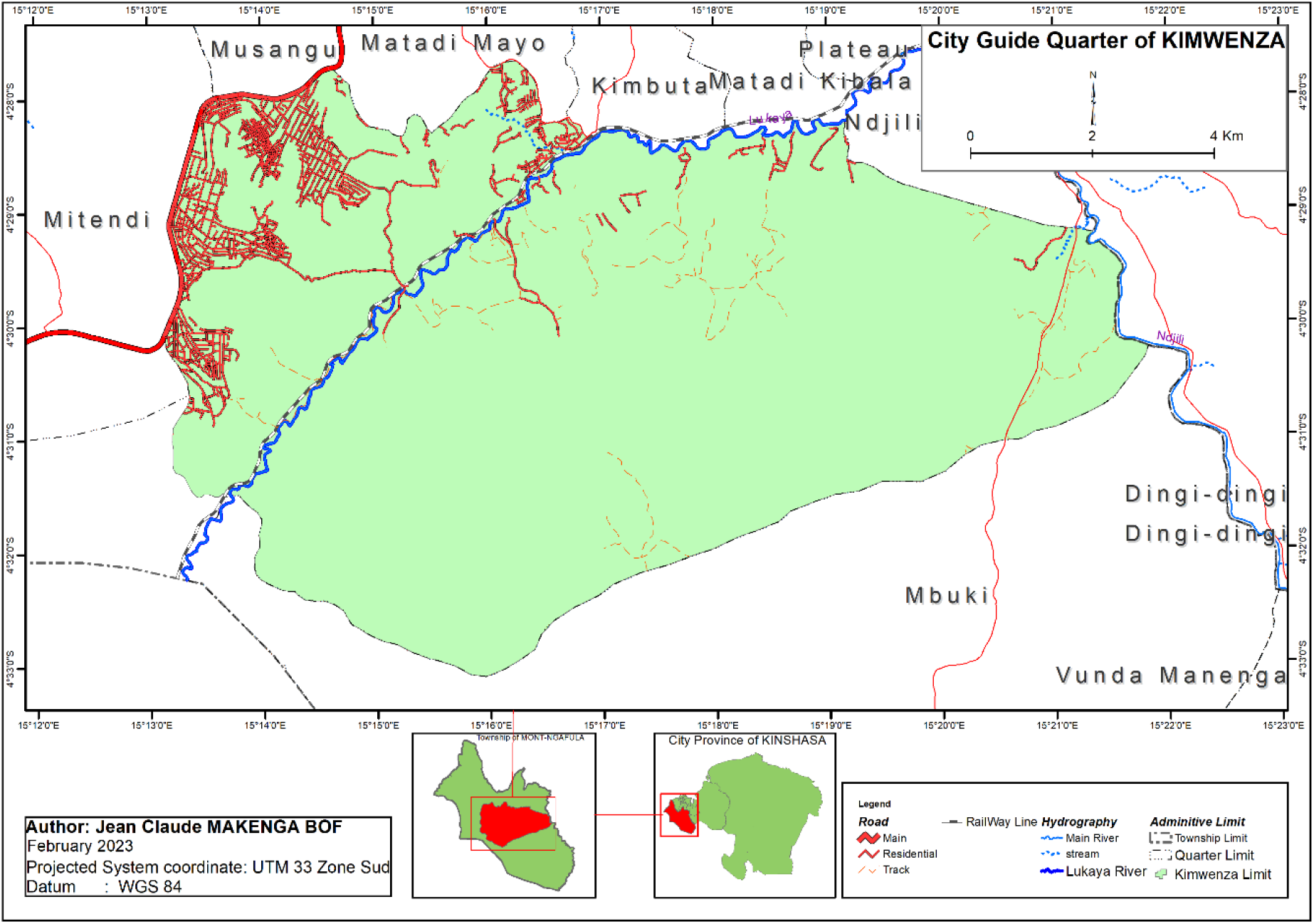
Map of the city-province of Kinshasa showing the site under study: Kimwenza Site in Mont-Ngafula 1, DRC, Democratic Republic of Congo.

### 2.2. Ethical clearance

The study was approved by the ethics committee of the Kinshasa School of Public Health (approval number ESP / CE / 139/2019). The data were collected following the ethical principles defined in the Declaration of Helsinki. Informed and written consent were obtained from the two black fly catchers used as human bait in this study. No financial incitive was given to blackfly catchers. However, they were provided medical examinations after participating in the study. The two blackfly catchers alternated after each hour to reduce their exposure times.

### 2.3. Capture of adult black flies

The study was conducted in a randomly chosen 24-week period, from August 1, 2019 to July 31, 2020. Blackfly catchers were trained on “human landing collection techniques” [16]. Berre’s method [11,17,18], slightly modified, were used to collect adult blackfly females. Blackflies were collected during five consecutive days per month, from 07:00 to 18:00 by 2 vector collectors working alternatively per hour using individual 6 ml polystyrene test tubes. (Greiner Ref: 148112). Collection tubes containing collected specimens were packed in wet cotton wool and kept in a well-shady place. Collected specimens were at the end of the day transferred in bottles containing alcohol. Each bottle was labelled with the catching point, date, and number of females preserved and all bottles were transferred to the bioecology laboratory of the school of Public Health of the University of Kinshasa (ESPK) for morphological examination and identification.

### 2.4. Identification by morphological criteria

Morphological identification was carried out at the bioecology laboratory of the ESPK using a VWR stereomicroscope model Stemi DV4. Blackflies were first anesthetized with chloroform vapor, and the morphological features were observed following the dichotomous keys proposed by Crosskey [19]. A few numbers of collected flies were dissected to determine their parity. The remaining flies were transferred into a vial (28 ml Universal Bottle, Ref: G751/01 Eurostar Scientific Ltd) filled with 80° alcohol. Each bottle was labelled (catching point, date, and number of females preserved) and shipped to the WHO/ESPEN laboratory in Ouagadougou, Burkina Faso for molecular analysis.

### 2.5. Determination of entomological indices

The biting rate transmission potential and infectivity rate [16, 20, 21], were used to determine entomological indices. The monthly biting rates (MBR) is the theoretical total number of bites that would be received by a person exposed for 11 hours a day near a river in one month. The Annual Biting Rate (ABR) is the sum of the 12 monthly biting rates. The Annual Transmission Potential (ATP) is the theoretical estimation of the total number of *O. volvulus* larvae that a person placed at the catching point under the conditions described above would receive in one year [16, 20, 22]. The vector infectivity rate refers to the number of infectious females (females with cephalic L3 parasites) per 1000 parous females, or the number of infectious flies per 2000 females analyzed (if the parturity rate is unknown, it is assumed to be 50% of the total population of biting females). In the present study, transmission and infectivity rates were determined using O-150 PCR technique, which is a diagnostic technique that aims to determine the level of infective-stage of *O. volvulus* larvae in the vector population as analyzed by polymerase chain reaction (PCR) technique based on amplification using *O. volvulus* specific DNA targeting probes O-150 repeat family sequence [23 - 27].

### 2.6. DNA extraction and purification

At the ESPEN Laboratory in Ouagadougou, *Simulium damnosum s*.*l*. flies were sorted out for the second time and pools were formed based on daily collection of blackflies, up to 100 blackflies maximum per pool. For each pool, the heads and bodies were separated by freezing and agitation and sifting by passage through a 25-mesh sieve, as previously described [17]. Only the head pools were processed. DNA extractions were carried out in sets of 20 samples each, with each set containing 18 fly pools and 2 sham extractions to ensure that the DNA extraction process remains free of contamination. The pools of heads were placed in a 1.5 ml microcentrifuge tube containing 200ul of 10mM Tris-HCl, 1mM EDTA (pH 8.0), and homogenized with a disposable plastic pastel. Homogenates were then subjected to proteinase K at final concentration of 2 mg/mL for 2 hours at 56°C. To digest the included proteins of *O. volvulus, d*ithiothreitol was added in samples to a final concentration of 20 mM and heated to 100°C for 30 minutes, to disrupt the di-Sulfide bonds abundant in the parasitic cuticle [28]. Boiling was followed by a series of three freeze-thaw cycles to release the DNA parasite into the media. The homogenates were centrifuged at 14000g for 5 minutes at 5°C and the supernatant placed into a new pre-labelled tube. The solutions were brought to a final concentration of 100 mM Tris-HCl (pH 7.5) 100 mM NaCl. Two oligonucleotide probes for the capture of *O. volvulus* DNA were used: One probe to capture the parasite genomic DNA (OVS2-biotin probe) and the other probe to capture the parasite mitochondrial DNA (Ovmito2353-biotin) [29 - 31]. A total of 5 ml of a 0.5 mM of each probe solution were added to each sample. Their DNA sequence are: for OVS2-biotin probe’s DNA sequence is as follow: (5’B-AATCTCAAAAAACGGGTACATA-3’, where B=biotin; while that of the Ovmito2353-biotin probe is: (5’ B-GTTTTAGGCTATTGGGCTG 3’, where B = biotin). The samples were then heated to 95°C for three minutes and allowed to cool slowly to room temperature. While the probe was annealing to the DNA in the solution, 1250 ml of Dynal M-280 streptavidin coated beads (Invitrogen) were aliquoted in 25 wells of a 96 well tissue culture plate. The plate was placed on a magnetic capture unit (DynaMag™-96 Bottom collection magnet, 96 well format, Invitrogen) and the beads collected for 2 minutes. The beads were then washed five times with 200 μl binding buffer (100 mM Tris-HCl (pH 7.5) 100 mM NaCl) per wash, resuspended in 50 ul of binding buffer and 10 μl was added to the 125 samples. The samples were incubated at 4°C overnight on a microplate Shaker & Incubator to permit the oligonucleotide-DNA hybrids to bind to the beads. The samples were placed in the magnetic separator for two minutes to capture the beads and the supernatant discarded. The beads were resuspended in 150 μl of binding buffer by pipetting, and the beads captured by placing the microplates in the magnetic separator for two minutes. The wash step was repeated five times. The beads were then resuspended in 20 μl of sterile water, heated to 80°C for 2 minutes and cooled rapidly on ice for two minutes. The beads were removed by placing the microplates in the magnetic capture unit, and the supernatant containing the purified DNA transferred to a new microplate and stored at -94°C.

### 2.7. O-150 PCR analysis

The purified DNA were tested for *O. volvulus* parasites by using a PCR assay specific for *O. volvulus*, as previously described [29, 32 - 35]. Seventy-four samples were tested in wells B1 to H2, Row A was reserved for 12 controls including 10 PCR negative controls (wells A1 to A10) and 2 positive controls (A11-A12). The positive control in A11 contained the minimum amount of positive control DNA consistently detected by PCR amplification conditions, as determined by an initial titration study. This control was performed to ensure that all reactions were running at maximum efficiency. The positive control in well A12 contained the same minimum amount of positive control DNA mixed with 2.5 μl of a DNA preparation from a pool that tested negative in a previous set of reactions. This control ensures that no inhibitors are present in the fly DNA preparations. 2.5 μl of the purified DNA containing genomic and mitochondrial DNA of *O. volvulus* were used as templates for the PCR amplifications carried out in a total volume of 50 μl containing 0.5 mmol/l of each O-150 primers : 5’-GATTYTTCCGRCGAANARCGC-3’ and 5’-B-GCNRTRTAAATNTGNAAATTC-3’, where B= biotin; N= A, G, C, or T; Y=C or T; and R=A or G. Reaction mixtures also contained 60 mM Tris-HCl, (pH 9.0), 15 mM (NH4)2SO4, 2 mM MgCl2, 0.2 mM each of dATP, dCTP, dGTP and dTTP, and 2.5 units of Taq polymerase. Cycling conditions consisted of five cycles of one minute at 94°C, two minutes at 37°C, and 30 seconds at 72°C, followed by 35 cycles of 30 seconds at 94°C, 30 seconds at 37°C, and 30 seconds at 72°C. and the final elongation step to have all amplicons with a double strand, at 72°C for six minutes.

The amplicons were detected by PCR enzyme-linked immunosorbent assay (ELISA), essentially as previously described [36]. In short, 5 μl of each PCR product was bound to a streptavidin-coated ELISA plate, and the DNA strands denatured by treatment with alkali. The bound PCR fragments were then hybridized to a fluorescein-labeled *O. volvulus*-specific oligonucleotide probe (OVS2: 5’-AATCTCAAAAAACGGGTACATA-FL-3’), and the bound probe detected with an alkaline phosphatase labeled anti-fluorescein antibody (fragment FA; Roche Diagnostics). Bound antibody was detected using the ELISA amplification reagent (BluePhos) kit from KPL (Gaithersburg, USA) following the manufacturer’s instructions. Color development was stopped by the addition of 100 μl AP stop solution, and the plates read in an ELISA plate reader set at 630 nm. Samples were scored positive if their optical density exceeded either the mean plus three standard deviations of ten negative control wells run in parallel or 0.1, whichever was greater.

### 2.8. Datal analysis

We calculated the confidence interval (95%) of annual biting rate (ABR), infectivity rate and annual transmission potential (ATP). Two-sample t-tests with unequal variances were used to compare the mean values of study parameters and the χ^2^ or Fisher’s exact tests were used to compare the proportions of parous females at Kimwenza site by months of capture. P <0.05 was considered significant. The data were analyzed using Excel ™ and SPSS 20 ™ Data Analysis Software.

## 3. Results

### 3.1. Vector Identification

All blackflies collected and examined belonged to the species *Simulium squamosum*.

### 3.2. Biting rates

A total of 12,217 black flies were captured during the study period, 22,380 Monthly variations in blackfly biting rates were very high. An analysis of MBR variations in relation to rainfall in the city of Kinshasa is presented in Figure 3.

**Figure 2:**
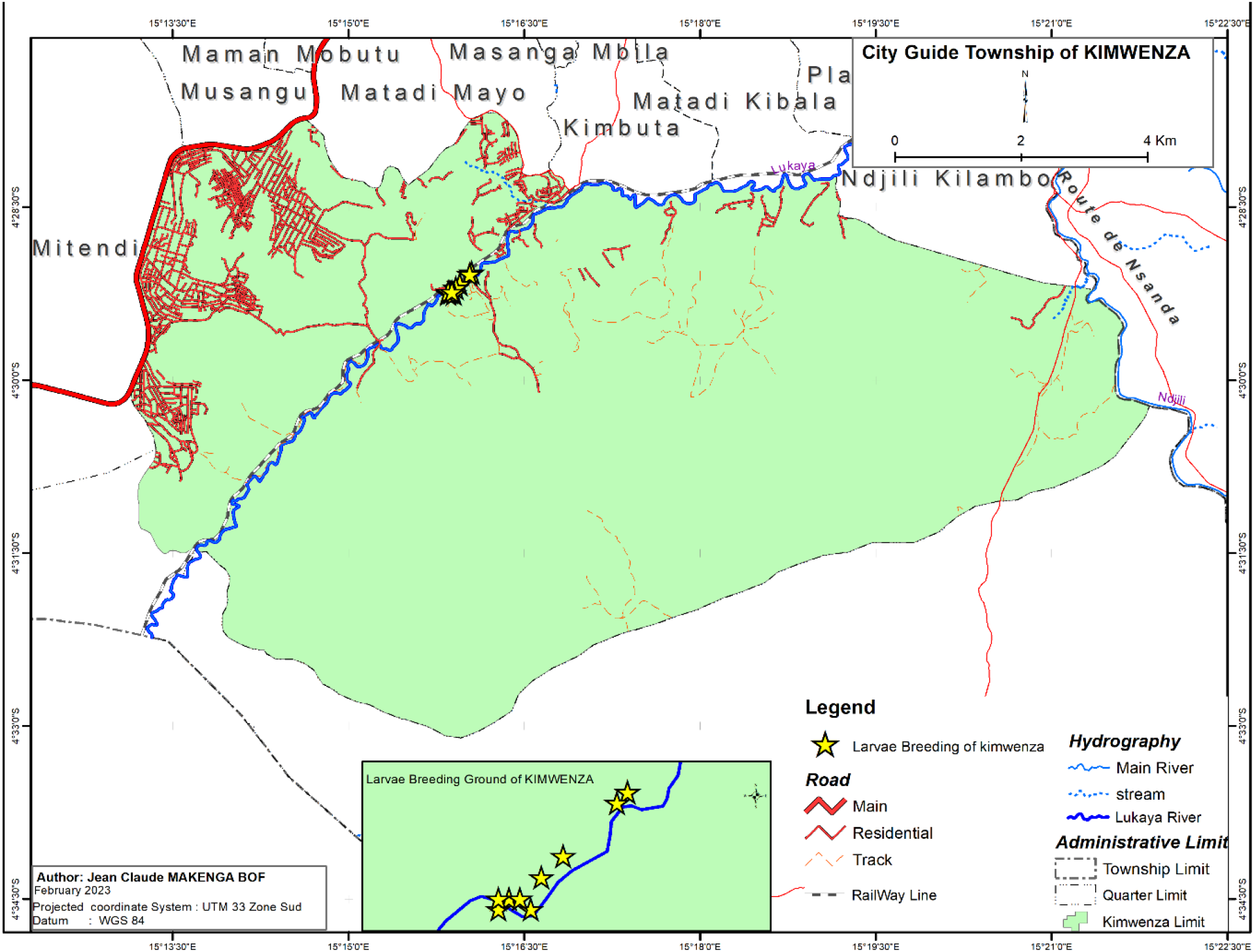
Geographical coordinates of the Kimwenza site in Mont-Ngafula DRC, Democratic Republic of Congo showing the collection site of black flies (“Simulia”) from August 2019 to July 2020.

**Figure 3:**
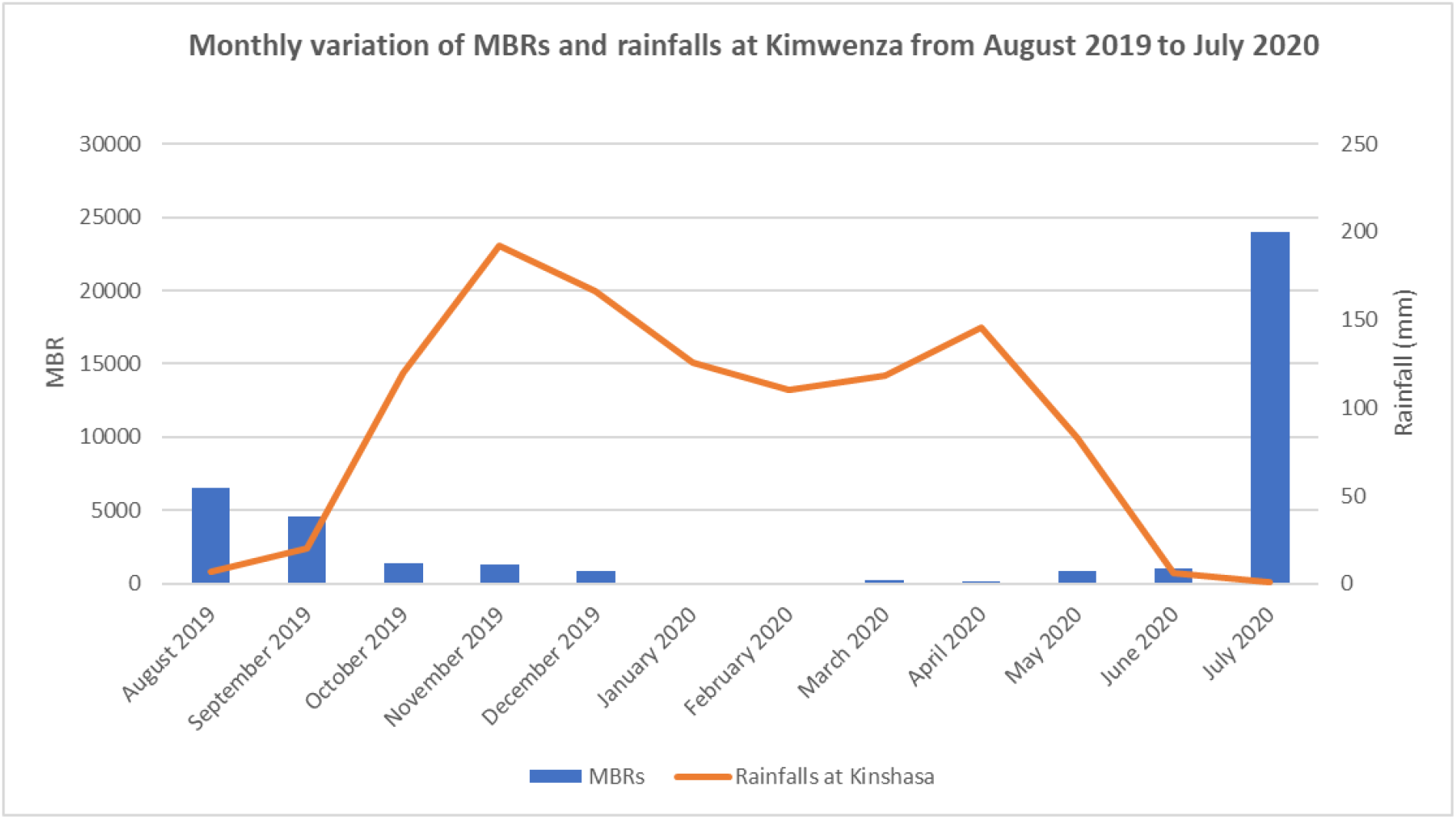
Monthly variation of Monthly Biting Rates (MBRs) and rainfalls at Kimwenza from August 2019 to July 2020.

During the rainy season, which begins in October and continues until May of the following year, blackfly productivity is very low. Identified supports were colonized by pre-imaginal stages of blackflies.

Two daily cycles of high biting were identified between 08:00 and 09:00 a.m. and 16:00 and 17:00. Low biting rates were observed between 11:00 a.m. and 13:00 (Figure 5).

**Figure 4:**
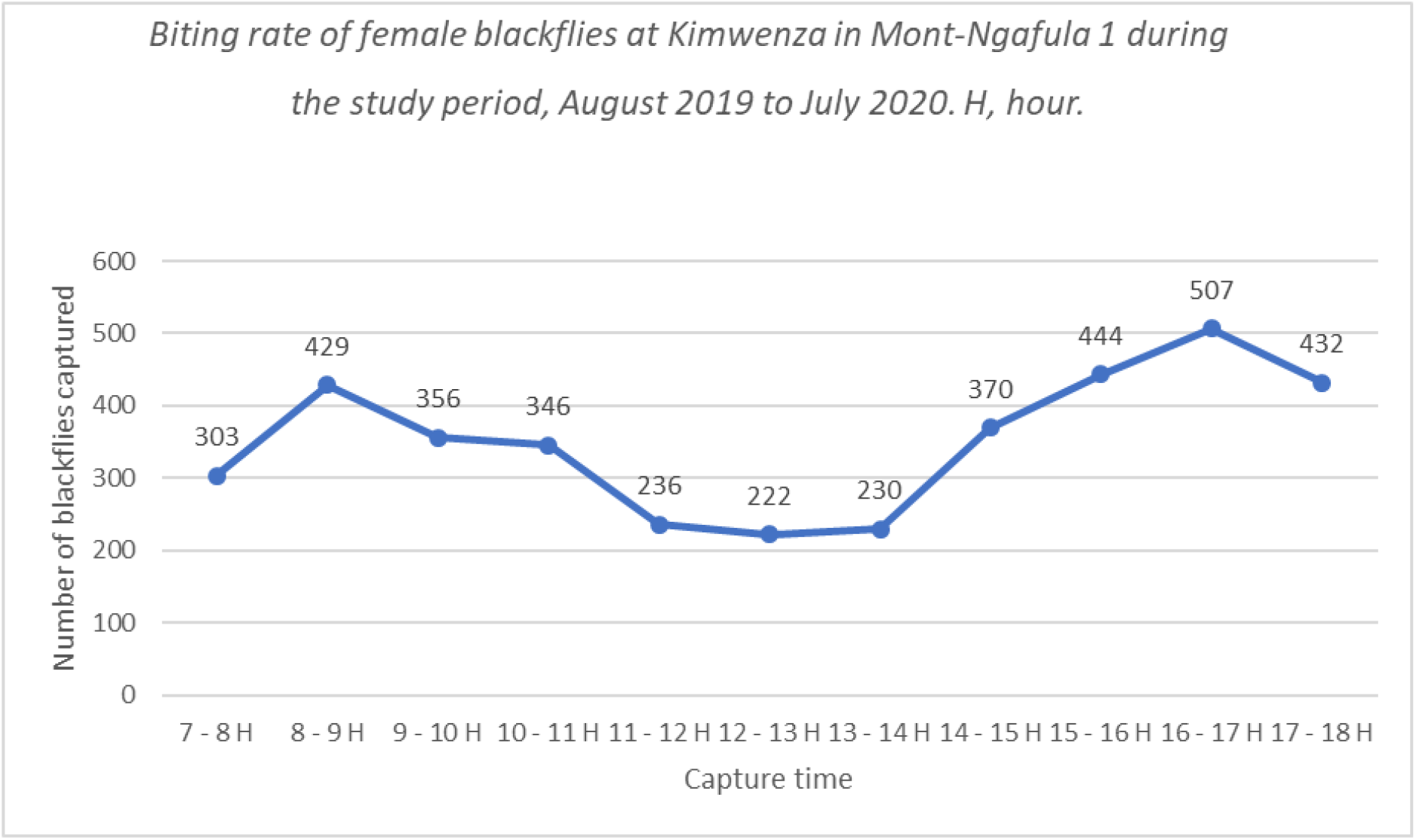
Biting rate of female blackflies at Kimwenza in Mont-Ngafula 1 during the study period, from August 2019 to July 2020. H, hour.

**Figure 5:**
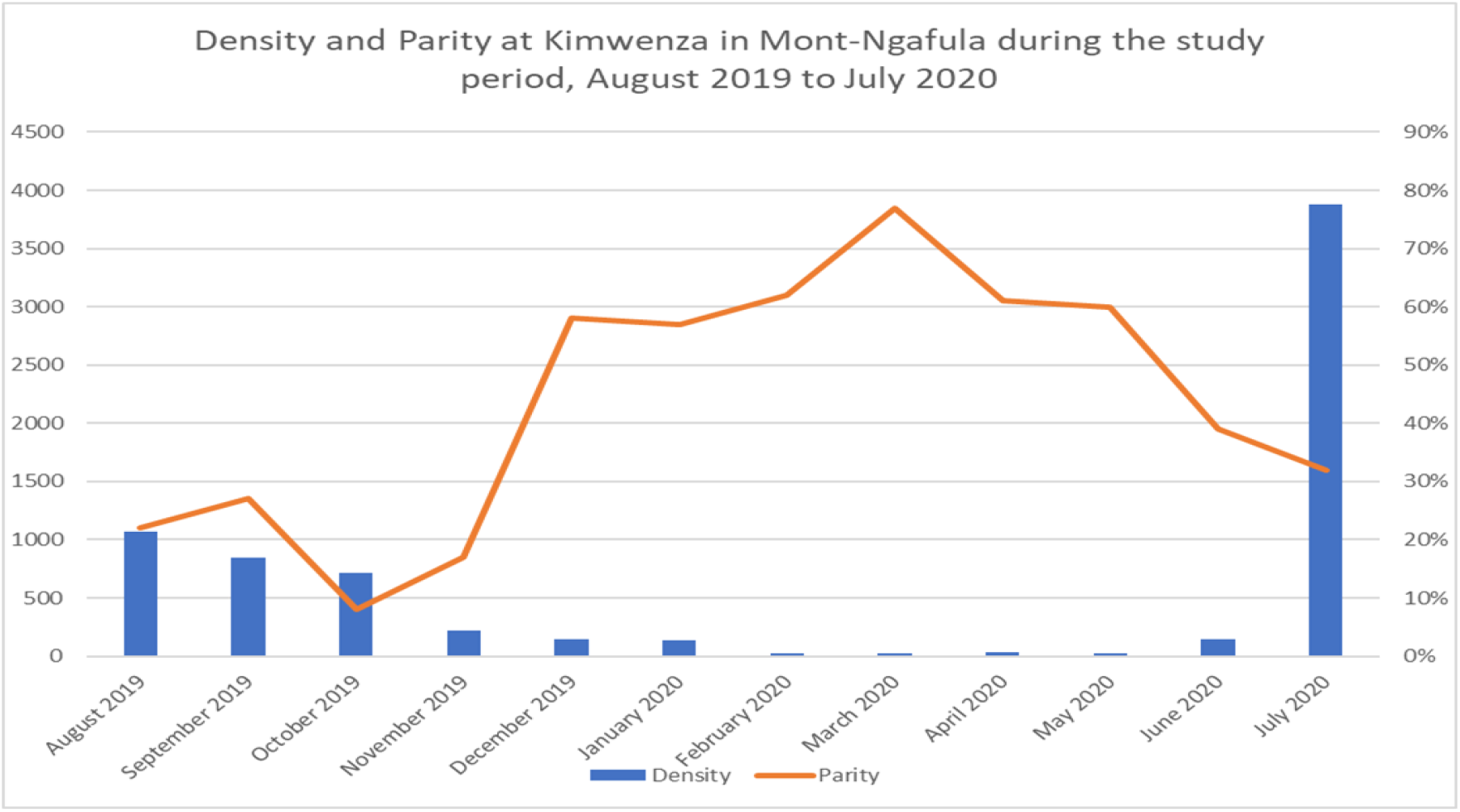
Variation of parous females at Kimwenza in Mont-Ngafula, from August 2019 to July 2020.

The ratio of parous to nulliparous allows us to estimate the survival rate of simulium, which is directly related to the risk of disease transmission. Dissection of 7,261 female flies resulted in an estimated parous females’ rate of 12% and nulliparous females’ rate of 88%. (Figure 5). The highest parities were obtained in February 2020 (62%), in March 2020 (77%), in April 2020 (61%) and in May 2020 (60%) (Figure 5).

### 3.3. Infectivity Rate and Annual Transmission Potential

The percentage of infected vectors, and the Annual Transmission Potential indices are presented in table 2. The blackfly infectivity rate is above the WHO recommended threshold of 0.05% (< 1/2000) in Kimwenza (P= 9.44%, 95% CI: 0.04 - 0.17)). The upper bound of the 95% confidence interval is above the recommended threshold meaning an ongoing transmission of *O. volvulus* infection. The calculated ATP was 21. Both the infectivity rate and ATP confirmed an ongoing transmission of *O. V* infection in Kimwenza.

**Table 1:** Number of blackflies collected and calculated biting rates (DBR, MBR and ABR) and percentage of parous and nulliparous females captured from August 2019 to July 2020 at the Kimwenza capture site in Mont Ngafula.

| Parameters | 2019 |  |  |  |  | 2020 |  |  |  |  |  |  | TOTAL |
| --- | --- | --- | --- | --- | --- | --- | --- | --- | --- | --- | --- | --- | --- |
|  | Aug | Sept | Oct | Nov | Dec | Jan | Feb | Mar | Apr | May | June | Jul |  |
| No. of capture days | 5 | 5 | 5 | 5 | 5 | 5 | 5 | 5 | 5 | 5 | 5 | 5 | 60 |
| No. of females caught (standardized & non-standardized) | 2,209 | 1,694 | 1,504 | 773 | 398 | 276 | 347 | 299 | 233 | 172 | 261 | 4,051 | 12,217 |
| No. of females caught using standardized catching techniques (HLC) and dissected | 1,072 | 847 | 712 | 223 | 144 | 138 | 21 | 26 | 36 | 25 | 142 | 3,875 | 7,261 |
| No. of females caught using unstandardized catching techniques | 1,137 | 847 | 792 | 550 | 254 | 138 | 326 | 273 | 197 | 147 | 119 | 176 | 4,956 |
| Daily Biting rate (DBR) | 214 | 169 | 142 | 45 | 29 | 28 | 4 | 5 | 7 | 5 | 28 | 775 | 1,452 |
| Monthly Biting Rate (MBR) | 3,323 | 2,541 | 2,207 | 669 | 446 | 428 | 61 | 81 | 108 | 78 | 426 | 12,013 | 22,380 |
| No. of parous females | 224 | 232 | 43 | 38 | 83 | 78 | 13 | 20 | 22 | 15 | 54 | 57 | 879 |
| Parous females (%) | 21 | 27 | 6.0 | 17 | 58 | 57 | 62 | 77 | 61 | 60 | 38 | 1.47 | 12 |
| No. of nulliparous females | 848 | 615 | 669 | 185 | 61 | 60 | 8 | 6 | 14 | 10 | 88 | 3,818 | 6,382 |
| Nulliparous females (%) | 79 | 73 | 94 | 83 | 42 | 43 | 38 | 23 | 39 | 40 | 62 | 99 | 88 |

**Table 2:**
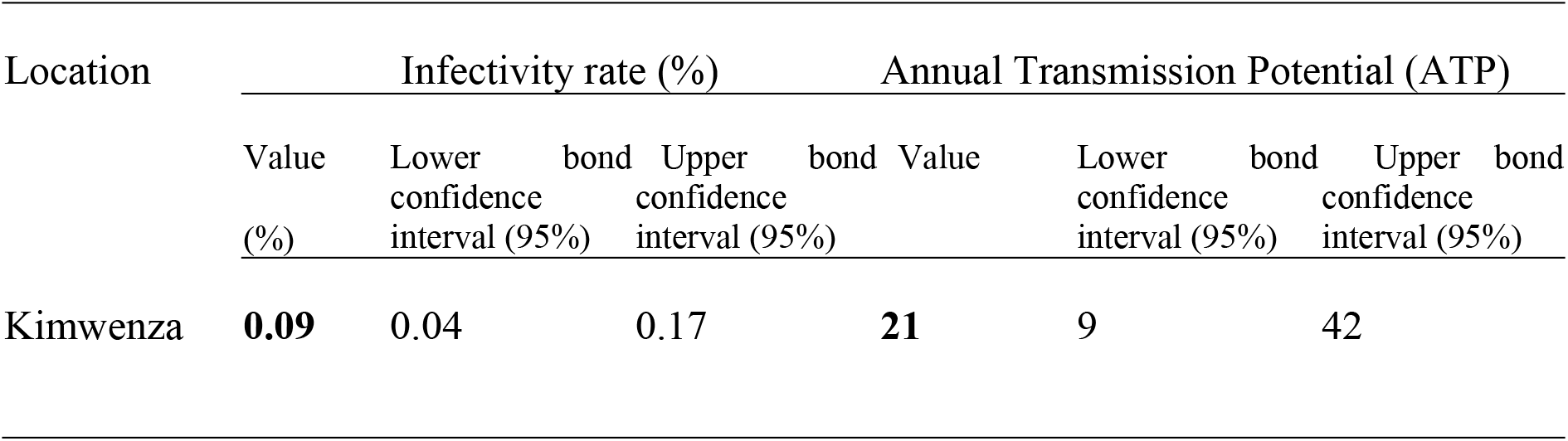
Transmission indexes at Kimwenza in Mont-Ngafula during the study period, August 2019 to July 2020.

### 3.4. Fixation supports of pre-imaginal forms in the capture sites

Different larval fixation supports were identified in the study area, from which pre-imaginal flies were collected: artificial waste (used cloths and plastic bags), aquatic plants (*Ledermaniella ledermannii, Tristicha trifaria, Pennisetum divisum, Phragmites mauritianus, Mimosa pigra, Cyperus distans, Ipomoea aquatica, Ludwigia adscendens, Echinochloa pyramidalis, Eichhornia crassipes, and Pistia stratiotes*), and plant remains (pieces of wood, tree branches, and palm leaves) wedged between rocks and trees.

These supports were found in abundance around the rapids of the Lukaya River especially in places frequented by humans from the surrounding areas. The pre-imaginal population was more prominent on *Ledermanniella ledermannii* (the main support)>artificial supports>plant remains.

## 4. Discussion

Morphological examination of the collected larvae and adult females revealed that they all belonged to the *S. squamosum* species of the *S. damnosum s*.*l*. complex. This was in alignment with results from previous surveys by Mansiangi (2012) [17] and Makenga (2015) [37] in Kinshasa, Henry *et al*. in 1984 in Kinsuka [11] and Hissette in Kinshasa in 1931 [38]. The pattern of MBR found in this study is in alignment with previous observations in Kinshasa [17, 37] and Brazzaville [49]. However, the ABRs is very high, with levels that could maintain a high transmission rate if there is a reservoir of parasites that escape annual ivermectin treatments. The threshold of 1000 bites/person/year by the *S. damnosum s*.*s*./*S. sirbanum* pair is the ABR value established by WHO below which there is no danger of human repopulation of onchocerciasis-free zones in the savannah zone of West Africa [50]. In ‘forest’ onchocerciasis regions where forest species of *S. damnosum s*.*l*. including *S. squamosum* are present, the tolerable thresholds of the ABR are far above the maximum values established in savanna zones. Considering the pattern of biting rates over the study period, it’s obvious that the good period for ivermectin mass campaigns should be between May to August. One would recommend organizing ivermectin mass campaigns during the high transmission season which in this case is between July and August of each year. This study showed blackfly infectivity rate (Upper bound of the 95% CI above 0.05%) and ATP (ATP =21) indices above the WHO recommended thresholds for the elimination of onchocerciasis [51].

In this study, estimates were calculated assuming an average of one L3 per infective fly. With this level of transmission, there is likely to be more than one L3 per fly, meaning that the ATP estimates are going to be too low. The assumption of one L3 per fly comes from OEPA’s experience of Demanou *et al*. 2003; Amuzu *et al*. 2010; Basáñez MG et al. 2009 and Duke et al. 1964 [44 - 47] but *S. damnosum* is a more efficient vector than is *Simulium metallicum, S. callidum*, and *S. ochraceum found in the Americas*. Also, in areas with good mass drug administration coverage, it can be hypothesized that most of the transmission will come from people that do not swallow ivermectin known as systematic non-compliers which constitute a reservoir of infection to systematic compliers. In these people, skin mf levels will be unchanged, and the number of L3 developing in a fly that gets a blood meal from the non-compliers is not going to differ from pre-control. Thus, in the perspective of elimination, if we cannot effectively reduce the densities of blackflies to break the chain of transmission, as was the case with vector control at onchocerciasis control program (OCP) in West Africa, it is even more opportune to choose the period of the MDA so as to keep microfilarodermia levels as low as possible, and prevent the dissemination of the parasite notwithstanding high biting rates.

This study shows a diurnal fly bite cycle with two peaks, the minor one between 08:00 and 09:00 and the major peak between 16:00 and 17:00 (Figure 5). Our results confirm the conclusions of Henry et al. (1984) who observed the same two peaks in the Kinsuka [11]. Mansiangi et al. also observed two peaks at Kinsuka: a major one between 16:00 and 17:00 and minor one between 09:00 and 10:00 [17]. Makenga et al. also observed two peaks: a major peak in the morning and minor peak around 17:00 [37]. This difference may have resulted from an increased fly activity when people were concentrated around the breeding sites, namely in the morning and at sunset, as in our case. Our results corroborate with those of Nascimento-Carvalho in Brazil or Homoxi, where the biting activity of female Simuliidae showed a bimodal pattern, with peaks in early morning (between 07:00 and 08:50.) and afternoon (between 16:00 and 17:50) [52]. In Thirei, the biting activity was evenly distributed throughout the day [53]. Black flies were captured simultaneously in Homoxi and Thirei using systematic methods to compare the traditional capture method using human bait (Human Landing Catch, HLC) with HLC protected by MosqTent® [52]. Comparing the anthropophilic profiles of black flies captured using MosqTent® and HLC, the percentage of species caught fluctuated seasonally in both Homoxi and Thirei, except between 11:00 and 11:50. in Homoxi and between 16:00 and 16:50. in Thirei, where the difference was based on the method of capture [52].

This study identified a diversity of larval breeding supports, such as waste, aquatic plants, and marine rocks, in and along the Lukaya rivers. Similar supports have been reported by Henry et al. (1984) [11], Mansiangi et al. (2014) [17], and Makenga et al. (2015) [37]. Considering the supports identified, this study confirms the breeding of *S. damnosum*; the flies lay their eggs in the river rapids, where the larvae hatch and develop into adults within eight to twelve days [54]. The pre-imaginal forms (eggs, larvae, and nymphs) are all aquatic and strongly rheophilic [54]. After hatching, the young larvae either remain attached to aquatic supports or drift with the current [54]. They feed with their rigid mandibular soles, which they use to randomly catch particles suspended in running water, including the nutrients they need [54].

## 5. Conclusion

This study showed high biting and transmission rates in the study site. These findings confirmed an ongoing transmission of *O*.*V* infection despite two decades of interrupted annual ivermectin mass distribution using CDTI strategy. To accelerate the interruption of the transmission of onchocerciasis and meet the 2030 WHO road map targets, there is an urgent need for the National Onchocerciasis Control Program stopping doing business as usual and change treatment strategies. Firstly, organizing ivermectin mass campaigns between June and July to be able to keep the microfilarodermia rate very low before the high transmission season. Secondly, the program should move from annual to biannual ivermectin mass distribution in the study area. Thirdly, the onchocercoasos program should couple Ivermectin mass distribution with slash and clear strategy to reduce both biting rate and nuisance and therefore accelerate the interruption of transmission.

## Data Availability

All data produced in the present work are contained in the manuscript

## 6. List of abbreviations

ABR: annual biting rate
APOC: African Programme for Onchocerciasis Control
ATP: annual transmission potential
CDTI: Community Directed Treatment with Ivermectin
DBR: daily biting rate
DRC: Democratic Republic of Congo
DTT: dichlorodiphenyltrichloroethane
ESPEN: Expanded Special Project for Elimination of Neglected Tropical Diseases
ESPK: Ecole de Santé Publique de Kinshasa
HLC: Human Landing Catch
ISTM: Institut Supérieur des Techniques Médicales
MBR: monthly biting rate
MDA: mass drug administration
MTP: monthly transmission potential
NPOC: National Program for Onchocerciasis Control
NS: Non-significant
NTDs: neglected tropical diseases
PNLMTN-CP: National Programme for Neglected Tropical Diseases Control
REMO: Rapid Epidemiological Mapping of Onchocerciasis
SD: Standard deviation
ULB: Université Libre de Bruxelles
UNIKIN: Université de Kinshasa
WHO: World Health Organization

## 6.1. Acknowledgments

The authors of this study sincerely thank the Ministry of Health of the DRC, the political and administrative leaders of Kinshasa, the authorities of the health zone of Mont-Ngafula I and Gombe, and all the participants who made this study possible.

## 6.2. Funding Statement

No funding sources.

## 6.3. Data Availability

The authors confirm that all data underlying the findings are fully available without restriction. All relevant data are within the paper and its Supporting Information files.

## 6.4. Competing interests

The authors declare that they have no competing interests.

## 6.5. Authors’ contributions

MBJC is the main author: he designed the study, participated to data collection and analysis. MBJC, MP, ZJ and YC participated to the writing of the manuscript, data analysis and marking of the final version. IF took part to data analysis and marking of the final version. DB, AA, SMS reviewed the manuscript. AA and SMS run the O-150 PCR. YC participated to the writing of the manuscript, methodology and read-through. All authors were involved in the preparation of the manuscript, edition and finalization of the version to be published and agreed to be accountable for all aspects related to the integrity of the work.

## Notes

### Competing Interest Statement

The authors have declared no competing interest.

### Funding Statement

This study did not receive any funding

### Author Declarations

Republique Democratique du Congo, Universite de Kinshasa, Ecole de Sante Publique, Comite d'Ethique, numero d'approbation ESP/CE/139/2019

